# Long-term longitudinal analysis of 4,187 participants reveals new insights into determinants of incident clonal hematopoiesis

**DOI:** 10.1101/2023.09.05.23295093

**Authors:** Md Mesbah Uddin, Seyedmohammad Saadatagah, Abhishek Niroula, Bing Yu, Whitney Hornsby, Shriienidhie Ganesh, Kim Lannery, Art Shuermans, Michael C. Honigberg, Alexander G. Bick, Peter Libby, Benjamin L. Ebert, Christie M. Ballantyne, Pradeep Natarajan

## Abstract

Clonal hematopoiesis (CH), characterized by blood cells predominantly originating from a single mutated hematopoietic stem cell, is linked to diverse aging-related diseases, including hematologic malignancy and atherosclerotic cardiovascular disease (ASCVD). While CH is common among older adults, the underlying factors driving its development are largely unknown. To address this, we performed whole-exome sequencing on 8,374 blood DNA samples collected from 4,187 Atherosclerosis Risk in Communities Study (ARIC) participants over a median follow-up of 21 years. During this period, 735 participants developed incident CH. We found that age at baseline, sex, and dyslipidemia significantly influence the incidence of CH, while ASCVD and other traditional risk factors for ASCVD did not exhibit such associations. Our study also revealed associations between germline genetic variants and incident CH, prioritizing genes in CH development. Our comprehensive longitudinal assessment yields novel insights into the factors contributing to incident CH in older adults.

## Introduction

Clonal hematopoiesis (CH) is a common aging-related phenomenon whereby blood cells are predominantly derived from a few hematopoietic stem and progenitor cells (HSPC) with acquired somatic mutation(s) in known leukemia driver genes that foster clonal expansion. CH with a variant allele fraction (VAF) ≥2% is termed clonal hematopoiesis of indeterminate potential (CHIP). The most frequently mutated genes in CH include epigenetic regulators *DNMT3A*, *TET2*, and *ASXL1* (DTA), splicing factors (SF) genes *SF3B1*, *SRSF2*, *U2AF1*; and DNA damage response (DDR) genes *TP53* and *PPM1D*; and *JAK2*^1^. CH is associated with many age-related conditions, including hematologic malignancy^2,3^, atherosclerotic cardiovascular disease (ASCVD)^4^, stroke^5^, chronic liver disease^6^, and heart failure^7^.

Clinical consequences of CH differ depending on driver genes, types of mutations, growth rate, and the size of the clones^8–11^. Though age is the strongest risk factor for the development of CH, most individuals do not develop CH. Additionally, genetic^1,12–15^ and environmental factors^16–21^ associate with increased odds of CH in cross-sectional analyses^22^, but their role in the incidence of CH has yet to be established firmly. Indeed, few studies^10,23,24^ have evaluated determinants of incident CH or progression of CH clones, defined as the new occurrence or expansion of clones (VAF≥2%) during long-term longitudinal follow-up. This study profiled incident CH in 4,187 middle-aged participants from the Atherosclerosis Risk in Communities Study (ARIC) over a median follow-up of 21 years to identify the determinants of incident CH in older age.

## Results

### CH at baseline visit

We investigated clonal hematopoiesis (CH) in a cohort of 10,871 participants from the ARIC Study baseline visits using whole-exome sequencing (WES) with the HiSeq 2000 platform (**Suppl. Table 1**). After excluding individuals with prevalent hematologic malignancy and those without WES data at a follow-up visit (**Suppl. Fig. 1**), our analysis focused on 4,187 study participants. **Table 1** and **Suppl. Table 1** presents the characteristics of these participants, among whom 2,478 (59.2%) were female, 951 (22.7%) were African American, and the mean (SD) age was 55.5 (5.5) years at the time of enrollment. A total of 576 CH clones with mutations in 51 driver genes were detected among 457 participants (**Suppl. Table 2**). Baseline CH prevalence was 10.9% (457/4187) at VAF≥2% and 3.8% (161/4,187) at VAF≥10% (**Fig. 1a**). Notably, in all age categories, most of participants with CH (383/457; 83.8%) carried a single mutated clone (**Fig. 1b**). The most frequently mutated genes included *DNMT3A*, *TET2*, and *ASXL1*, with median VAF ranging from 6.6 to 15.3% (**Fig. 1c-d**). Our study provides comprehensive insights into the prevalence and characteristics of CH in middle-aged participants.

**Fig. 1.**
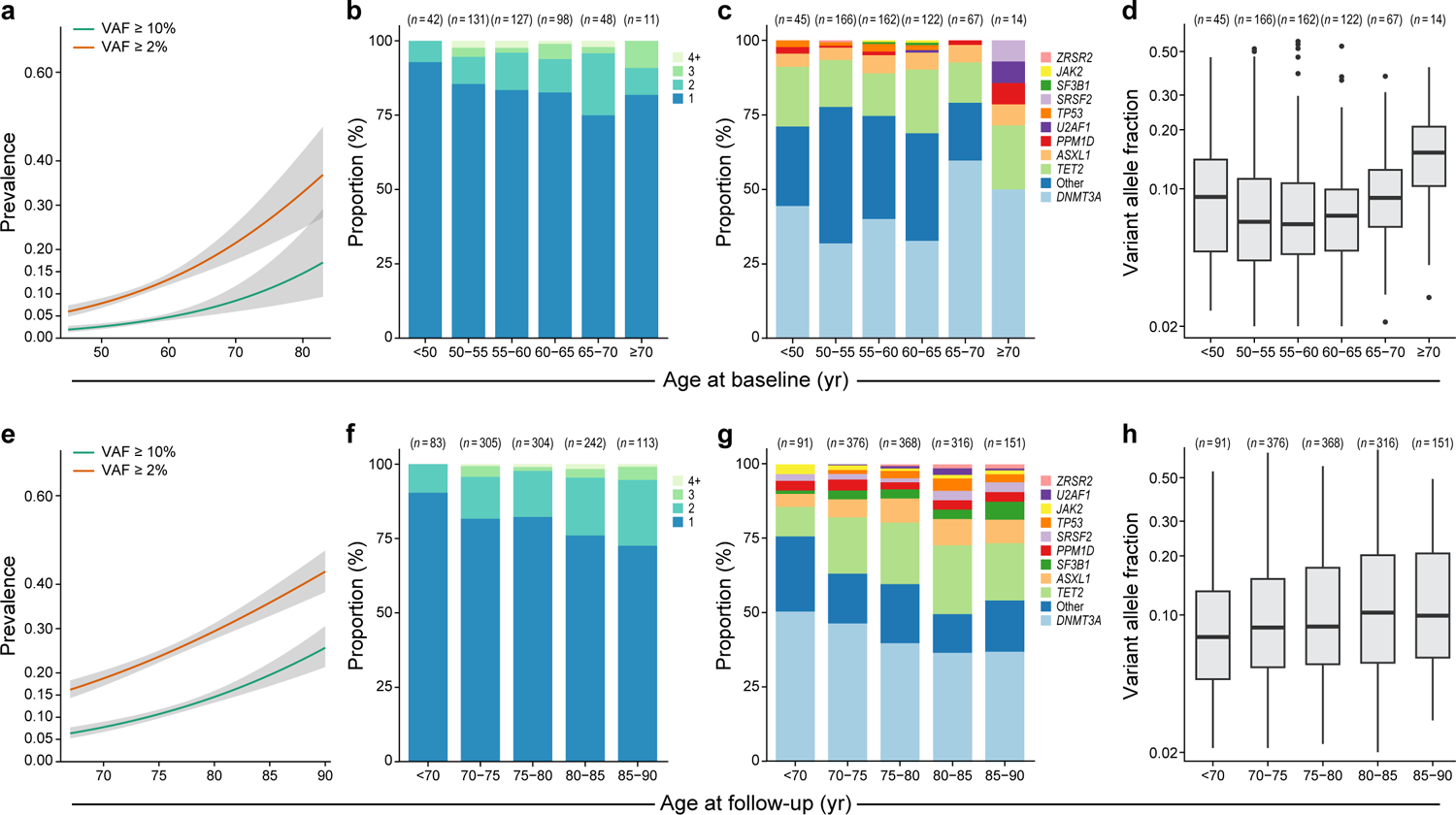
Distributions of clonal hematopoiesis (CH) at baseline (a-d) and follow-up (e-h) visits. CH prevalence increases with age, reaching approximately 30% when individuals reach 80 years (a, e). Most individuals with CH typically carry a single clone (b, f). In the earlier decades of life (under 70 years), many individuals carry mutations in *DNMT3A* or other less commonly mutated CH genes (c, g). However, individuals aged 70 and older show a higher incidence of CH involving genes such as *TET2*, *ASXL1*, and splicing factors (c, g), and the size of these clones tends to be relatively larger in later years (d, h). VAF: variant allele fraction.

**Table 1.** Characteristics of the participants with or without prevalent clonal hematopoiesis (CH) in the Atherosclerosis Risk in Communities Study.

| Characteristics |  | Without Prevalent CH | With Prevalent CH | *P* |
| --- | --- | --- | --- | --- |
| N |  | 3,730 | 457 | - |
| Age, mean (SD), y |  | 55.3 (5.4) | 57.2 (6.0) | 2.7E-10 |
| Sex | Female, n (%) | 2,226 (59.7) | 252 (55.1) | 0.07* |
|  | Male, n (%) | 1,504 (40.3) | 205 (44.9) |  |
| Ancestry | African American (AA) | 865 (23.2) | 86 (18.8) | 0.04* |
|  | European American (EA) | 2,865 (76.8) | 371 (81.2) |  |
| Ever Smoker, n (%) |  | 2,004 (53.7) | 265 (58.0) | 0.08 |
| BMI, mean (SD), kg/m<sup>2</sup> |  | 27.8 (5.1) | 27.6 (5.2) | 0.40 |
| SBP, mean (SD), mmHg |  | 118.3 (16.3) | 119.5 (16.6) | 0.14 |
| DBP, mean (SD), mmHg |  | 72.5 (9.8) | 72 (9.7) | 0.35 |
| Cholesterol medication, n (%) |  | 197 (5.3) | 34 (7.5) | 0.09 |
| Hypertension, n (%) |  | 1,056 (28.4) | 138 (30.2) | 0.42 |
| T2D, n (%) |  | 305 (8.2) | 44 (9.7) | 0.31 |
| CHD, n (%) |  | 116 (3.1) | 11 (2.4) | 0.36 |
| IS, n (%) |  | 38 (1.0) | 4 (0.9) | 0.78 |
| HF, n (%) |  | 103 (2.8) | 10 (2.2) | 0.45 |
| Total cholesterol, mean (SD), mg/dl |  | 207.9 (36.3) | 209.7 (38.1) | 0.35 |
| LDL-C, mean (SD), mg/dl |  | 130.5 (34.4) | 131.3 (30) | 0.65 |
| HDL-C, mean (SD), mg/dl |  | 52 (16.9) | 51.7 (17.9) | 0.67 |
| Triglycerides, median (IQR), mg/dl |  | 109.0 (75) | 119.0 (76) | 0.02<sup>#</sup> |
| Follow-up, mean (range), y |  | 20.3 (5-26) | 20.1 (6-27) | 0.07 |

*P*-values are from a two-sided *t*-test comparing baseline samples with and without prevalent CH. \**P* from Pearson's *Chi-squared* test. <sup>#</sup>*P* from a two-sided Wilcoxon rank sum test. BMI: Body mass index; CH: clonal hematopoiesis; CHD: Coronary heart disease; DBP: Diastolic blood pressure; HDL-C: high-density lipoprotein cholesterol; HF: Heart failure; IQR: interquartile range; IS: ischemic stroke; LDL-C: low-density lipoprotein cholesterol; SBP: Systolic blood pressure; SD: standard deviation; T2D: Type 2 Diabetes.

### CH at the follow-up visit

We further ascertained CH at a later visit, with a median follow-up duration of 21 years (range=5-27; mean=20.3; SD=2.03). The mean (SD) age of participants at follow-up blood draw was 75.8 (5.2) years. For this analysis, we performed WES on 4,187 samples using the NovaSeq 6000 platform and identified 1,302 CH clones at VAF≥2% in 50 driver genes among 1,047 participants (**Suppl. Table 3**). The prevalence of CH showed an age-dependent increase, reaching an overall prevalence of 25.0% (1047/4187) and 11.8% (496/4187) at VAF≥2% and ≥10%, respectively (**Fig. 1e-f**). With advancing age, we observed a shift in the proportions of individuals carrying specific CH subtypes. While the prevalence of *DNMT3A* CH and mutations in other less-frequently mutated genes decreased, mutations in *TET2*, *ASXL1,* and splicing factor genes increased (**Fig. 1g**). Additionally, clone sizes tended to increase with advancing age, with a median VAF ranging from 7.7 to 10.2% (**Fig. 1h**). The shifting patterns and increasing clone sizes of CH subtypes in older adults show a dynamic nature of CH over time.

### Concordance of CH calls from the HiSeq vs NovaSeq sequencing platforms

Here, baseline and follow-up visits were sequenced using two different sequencing platforms, HiSeq 2000 and NovaSeq 6000. We re-sequenced 786 samples from the same baseline visit using NovaSeq to assess the systematic difference in estimated VAF and the concordance of CH ascertainment between the two sequencing platforms. Concordance estimates for CH clones detected “yes/no” by the two sequencing platforms were 83% (654/786) and 93% (731/786) at VAF≥2% and VAF≥10%, respectively. We also observed a strong correlation (Pearson’s r=0.80) between the VAF estimates from these two platforms (**Suppl. Fig. 2**). As the VAF did not correlate perfectly, we only focused on incident CH, defined as a CH clone at VAF≥2% detected at the follow-up visit only without any prevalent CH clone at baseline, for subsequent analyses.

### Incident CH in the ARIC Study

We identified 3,730 participants without prevalent CH, of which 59.7% (2,226/3,730) were female, 23.2% (865/3,730) were African American, and 53.7% (2,004/3,730) had a history of smoking (**Table 1**). A total of 735 (19.7%) participants developed incident CH (VAF≥2%) during the follow-up, of which 37% (272/735) had large clones (VAF≥10%). Individuals with incident CH were relatively older at the baseline visit (median age of 56 vs 54 years; Wilcoxon rank sum test *P*=2.4E-6). Here, 876 incident clones were detected in 735 participants, where the majority (615/735; ∼84%) acquired a single clone during follow-up (**Fig 2a**). Most incident CH mutations occurred in DTA (*DNMT3A, TET2,* or *ASXL1*), followed by SF (*SF3B1, SRSF2, U2AF1,* or *ZRSR2*) and DDR (*PPM1D* or *TP53*) genes, and large clones (VAF≥10%) in *ASXL1*, *SF3B1, JAK2, ZNF318, U2AF1*, and *ZRSR2* (**Fig 2bc**). CH incidence increased with advancing age, where >23% of participants older than 75 years acquired incident CH (**Fig. 2d**).

**Fig. 2.**
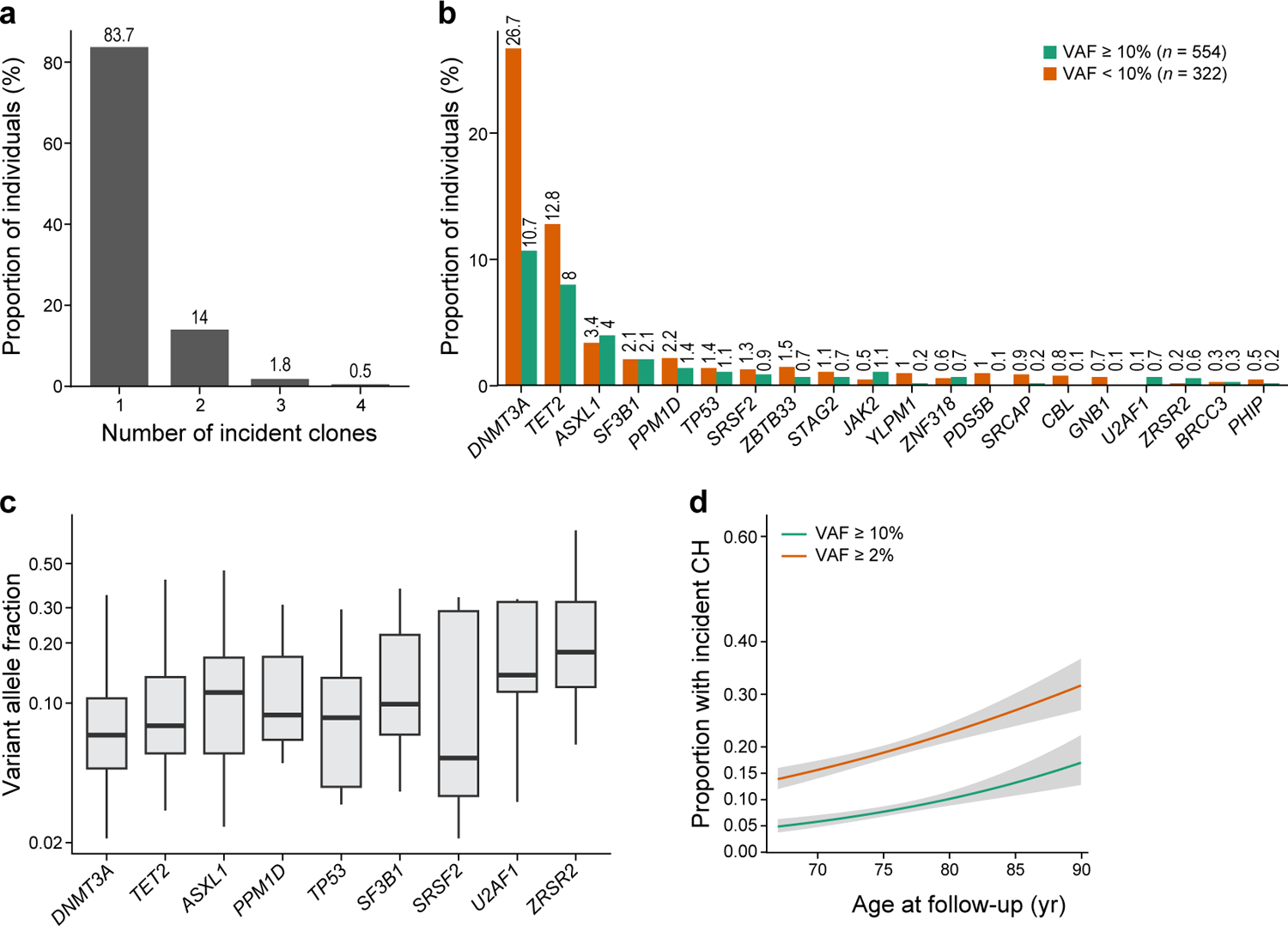
Distribution of incident clonal hematopoiesis (CH) among older adults. (a) The vast majority (over 83%) of individuals with incident CH carry a single clone, (b) with approximately 37% of the clones exhibiting an expanded state (VAF>=10%). (b) *DNMT3A* and *TET2* show higher proportions of smaller incident clones (VAF between 2% and 10%), while *ASXL1, JAK2, U2AF1*, and *ZRSR2* display expanded clones. (c) The median clone size exceeds 10% for *ASXL1, SF3B1, U2AF1*, and *ZRSR2* CH. (d) Like CH prevalence (Fig 1), CH incidence increases with age. VAF: variant allele fraction.

### Clinical predictors of incident CH

First, we performed univariable logistic regression to examine the associations of baseline risk factors such as age, sex, ancestry, body mass index (BMI), high-density lipoprotein cholesterol (HDL-C), non-HDL-C, history of smoking, hypertension, ASCVD, and T2D with incident CH (**Suppl. Fig. 3**). In univariable analyses, we observed significant associations (*P*<0.0025, considering 20 independent tests at a 5% level of significance) of age at baseline, male sex, and European ancestry with incident CH categories (**Suppl. Fig. 3**). Age was significantly associated with higher incidence of overall CH, *TET2*, and SF mutations (1.03≤ odds ratio (OR) ≤1.09; 5.4E-06≤ *P* ≤3.2E-4), and nominally associated (0.0025<*P*<0.05) with incidence of *ASXL1* and DDR mutations (**Suppl. Fig. 3**). However, no association was found between age and incident *DNMT3A* mutations. Male sex was significantly associated with a higher incidence of overall CH, *ASXL1*, and SF (1.32≤ OR ≤2.82; 6.9E-5≤ *P* ≤9E-4) and nominally associated with higher incidence of DDR mutations (**Suppl. Fig. 3**). European ancestry was significantly associated with lower incidence of *DNMT3A* (OR=0.67; 95% confidence interval (CI)=0.52-0.86; *P*=1.8E-3), and nominally associated with higher incidence of SF mutations (**Suppl. Fig. 3**). Additionally, there were nominal nonsignificant (0.0025<*P*<0.05) associations between smoking status (never vs. ever) and incident SF, BMI (inverse-rank normalized) and incident *DNMT3A*, history of ASCVD and incident *DNMT3A*, and non-HDL-C level (inverse-rank normalized) and incident *TET2* (Suppl. Fig. 3).

Next, we performed multivariable-adjusted logistic regression analyses of incident CH categories vs. baseline risk factors, including age, sex, race, BMI, HDL-C, non-HDL-C, history of smoking, hypertension, ASCVD, and T2D (**Fig. 3**, and **Suppl. Fig. 4**). Fully adjusted regression models included baseline risk factors and covariates such as age, sex, race, BMI, HDL-C, non-HDL-C, cholesterol-lowering medication use, history of smoking, hypertension, ASCVD, T2D, baseline visits, and visit center. Age was independently associated with a higher incidence of overall CH, as well as gene-specific CH subtypes (1.04≤ OR ≤1.10; 2.0E-7≤ *P* ≤7.4E-5), with higher odds for splicing factors genes (**Fig. 3** and **Suppl. Fig. 4**). Male sex was nominally associated with a higher incidence of overall and DDR CH, and significantly associated with a higher incidence of *ASXL1* and SF CH (1.30≤ OR ≤2.79; 7.5e-4≤ *P* <0.05). European ancestry was nominally associated with a lower incidence of CH in *DNMT3A* and a higher incidence of CH in DDR. Interestingly, no significant association was observed between BMI, HDL-C, non-HDL-C, history of smoking, hypertension, T2D, and ASCVD, and incident CH categories (**Fig. 3** and **Suppl. Fig. 4**). However, we observed a nominal non-significant association of higher BMI with reduced incident *DNMT3A*, and history of ASCVD with increased incident *DNMT3A* but reduced incident *TET2* (**Fig. 3**).

**Fig. 3.**
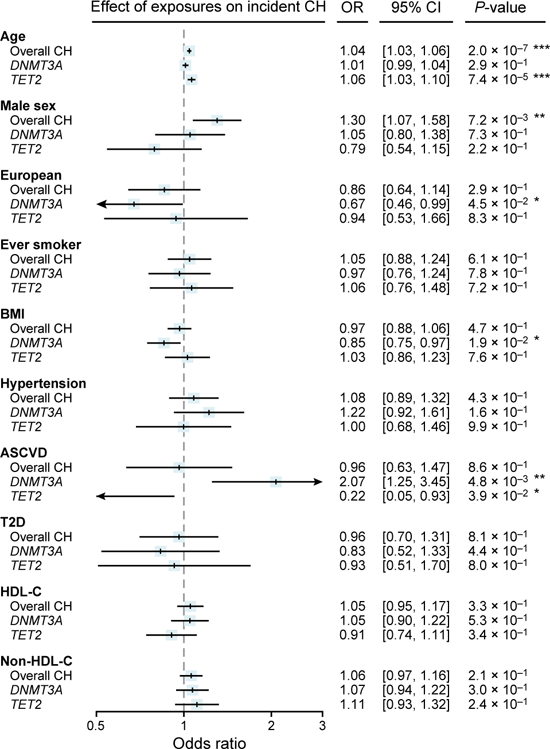
Association of clinical cardiovascular risk factors with incident clonal hematopoiesis (CH). Logistic regression analysis examined the association between incident CH categories and baseline cardiovascular risk factors. The adjusted model accounted for age, sex, race, body mass index (BMI), high-density lipoprotein cholesterol (HDL-C), non-HDL-C, cholesterol medication usage, smoking history, hypertension, atherosclerotic cardiovascular disease (ASCVD, including coronary heart disease and/or ischemic stroke), type 2 diabetes (T2D), and batch effects. Inverse rank normalization was performed before the analysis to account for potential variations in BMI, HDL-C, and non-HDL-C distribution. The results indicate that age is independently and significantly associated with a higher incidence of CH. Additionally, nominal non-significant associations are observed for male sex, European ancestry, BMI, and history of ASCVD with incident CH categories. ***: *P*<0.0025 (0.05/20); **: *P*<0.01; *: *P*<0.05.

In secondary analyses we tested the associations of smoking categories (never vs. former or current smokers), BMI categories (BMI <25 vs. 25-30 or >30 kg/m^2^), triglyceride to HDL-C (TG/HDL-C) ratio, dyslipidemia, male sex by smoking status (never vs. ever) with incident CH categories in fully adjusted models (**Suppl. Fig 5-6**). There were no significant associations between smoking status (never vs. former or current smoker) or BMI categories and incident CH categories (**Suppl. Fig. 5**). However, we observed a nominal non-significant association between current smoking status and incident CH in splicing factor genes, and between BMI >30 kg/m^2^ and incident *ASXL1* (**Suppl. Fig. 5**).

No significant associations were observed between TG/HDL-C ratio and incident CH (**Suppl. Fig. 6a**), although dyslipidemia did significantly associate with increased odds of incident *TET2* (OR=2.51; *P*=2.5E-3; **Suppl. Fig. 6b**). A nominal association was observed between dyslipidemia and increased odds of incident *ASXL1* (**Suppl. Fig. 6b**). We also tested interactions between sex and smoking history vs. incident CH categories in exploratory analyses. Fully adjusted models did not reveal significant interactions between sex and smoking history on incident CH categories. Nevertheless, there was nominal interaction between sex and smoking history: we observed lower odds of incident *DNMT3A* in the male sex by ever-smoker status (OR=0.48; *P*=0.0036) (**Suppl. Fig. 6c**).

### Shared genetic predisposition in prevalent and incident CH

We separately assessed the association of an independently-derived prevalent CH polygenic risk score (PRS), consisting of 21 conditionally independent and genome-wide significant (*P*<5E-8; **Suppl. Table 4**) variants^13^, with incident CH categories in African American (AA) and European American (EA) participants, followed by inverse-variance weighted fixed-effect meta-analysis (**Fig. 4ab**). We found that per SD increase in genetic liability to prevalent CH was significantly associated with incident CH (OR=1.23; 95% CI 1.12-1.35; *P*=8.9E-6). Furthermore, genetic liability to prevalent CH was associated with incident *DNMT3A* (OR=1.28; 95% CI 1.12-1.46), *TET2* (OR=1.30; 95% CI=1.09-1.55), and *ASXL1* (OR=1.66; 95% CI=1.26-2.17) CH (**Fig. 4b**).

**Fig. 4.**
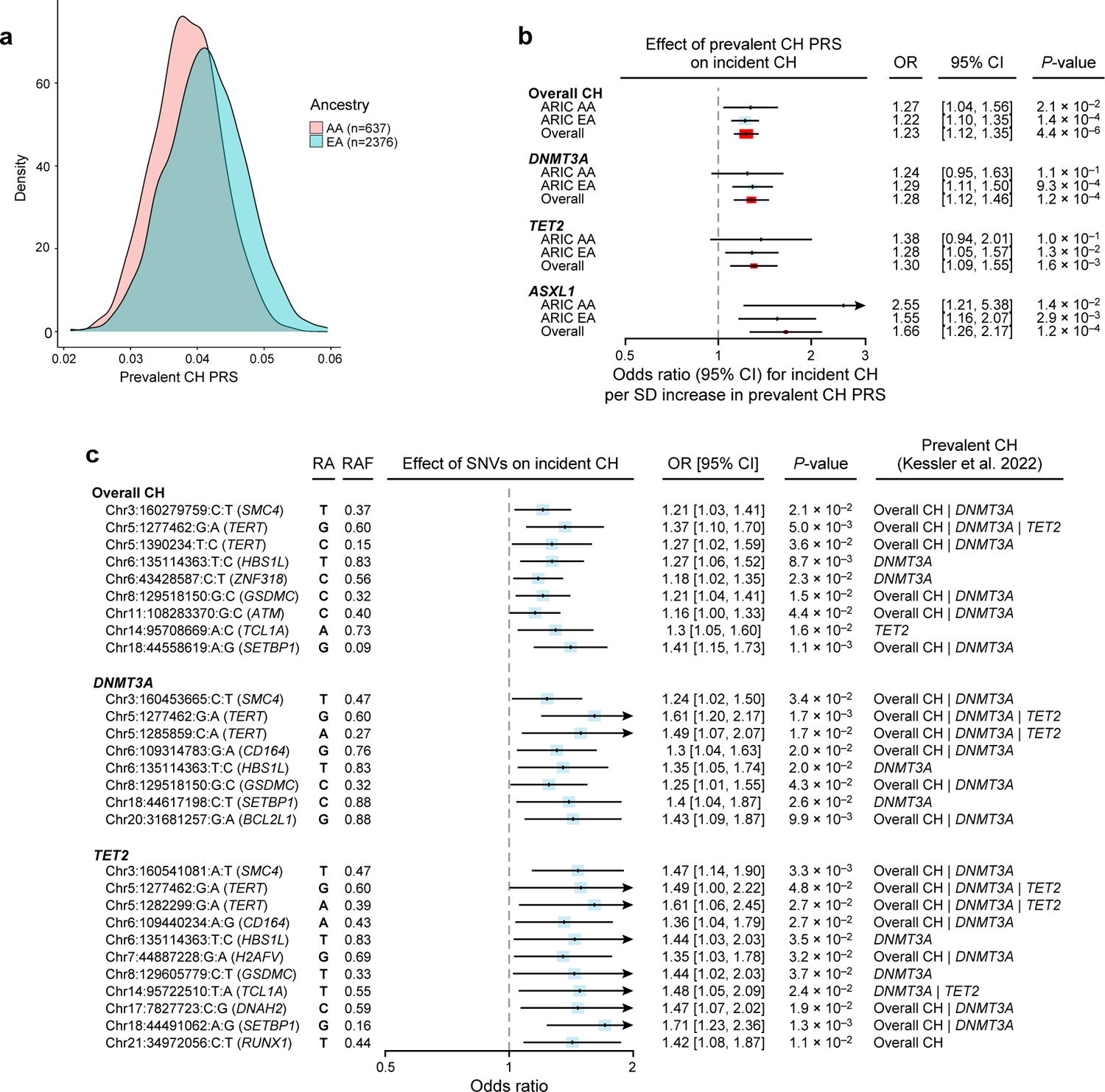
Association of prevalent clonal hematopoiesis (CH) with incident CH: (a-b) polygenic risk score (PRS) and (b) germline variants analysis. a) Distribution of PRS in ARIC AA and EA participants. b) Ancestry-stratified PRS was calculated using 21 independent variants (*P*<5e-8; Supplementary Table 4) from Kessler, et al. ^13^. Logistic regression was performed, adjusting for age, sex, smoking status, top five principal components of ancestry, and batch effect. The results show a strong association between prevalent CH PRS and incident CH. c) Single-variant association adjusted for age, age^2^, sex, top ten principal component of ancestry, and batch effect, followed by multi-ancestry meta-analysis was conducted to investigate the association of genome-wide significant (*P*<5e-8) prevalent CH-associated variants^13^ and incident CH at a significant level of *P*<0.05. This figure presents the lead variant(s) from each locus, and the full list is available in Supplementary Tables 5-7. Odds ratios are provided for the risk-increasing alleles. Several germline variants previously associated with prevalent CH were found to be associated with incident CH, indicating a shared genetic basis between prevalent and incident CH. ARIC AA: the Atherosclerosis Risk in Communities Study African American; ARIC EA: European American; RA: risk allele; RAF: risk allele frequency; SNV: single nucleotide variant.

Next, to test the associations of genome-wide significant (*P*<5E-8) variants for prevalent CH with risk of incident CH, we performed targeted single variant associations in AA and EA participants from ARIC. We tested known loci associated with prevalent CH derived in the UK Biobank by Kessler, et al. ^13^. For matching variants with minor allele frequency≥1%, we performed ancestry-stratified single-variant associations for incident CH, *DNMT3A*, and *TET2* in the ARIC study, followed by a multi-ancestry inverse-variance weighted fixed-effect meta-analysis. A *P* threshold of <0.05 was considered statistically significant. At this threshold, we reported several prevalent CH loci associated with incident CH (**Fig. 4c** and **Suppl. Tables 5-7**). Notably, the risk alleles in *SMC4, TERT, HBS1L, ZNF318, GSDMC, ATM, TCL1A*, and *SETBP1* loci were associated with a higher incidence of overall CH (1.16≤ OR ≤1.41; 0.0011≤ *P* ≤0.044; **Fig. 4c and Suppl. Table 5**). Risk alleles in *SMC4*, *TERT*, *CD164*, *HBS1L*, *GSDMC*, *SETBP1*, and *BCL2L1* loci were associated with higher incidence of *DNMT3A* CH (1.24≤ OR ≤1.61; 0.0017≤ *P* ≤0.043; **Fig. 4c** and **Suppl. Table 6**). The risk alleles in *SMC4, TERT, CD164, HBS1L, H2AFV, GSDMC, TCL1A, DNAH2, SETBP1,* and *RUNX1* loci were associated with higher incidence of *TET2* CH (1.35≤ OR ≤1.71; 0.0013≤ *P* ≤0.048; **Fig. 4c and Suppl. Table 7**).

## Discussions

We conducted one of the largest long-term longitudinal whole-exome sequencing studies on clonal hematopoiesis involving 4,187 healthy participants from the ARIC study with a median follow-up of 21 years. We identified CH (VAF≥2%) in 11% of the participants at baseline and 25% at the follow-up visits. Consistent with previous studies^1,4^, we showed that the prevalence of CH increases with advancing age.

We observed incident CH in approximately 17% of participants below 70 years, and this proportion increased to around 30% in individuals above 85 years of age among those without CH in middle age. These results reinforce the substantial impact of age on the acquisition of CH. Furthermore, our study revealed an interesting pattern of decreasing diversity in mutated CH genes with advancing age. Specifically, we noted an increase in incident clones in genes such as *TET2*, *ASXL1*, and splicing factor genes (*SF3B1*, *U2AF1*, *SRSF2*, *ZRSR2*), as well as *PPM1D* and *TP53*. These findings corroborate and extend recent reports highlighting reduced clonal diversity, faster expansion, and the acquisition of new mutations in these genes as individuals age^10,25,26^.

Previous cross-sectional studies reported associations of age, smoking, T2D, and BMI with prevalence of CH^2,3,16,27^. Interestingly, the impact of age on CH subtypes varied, with stronger associations observed for splicing factor genes, *TET2*, and *ASXL1* CH, but not for *DNMT3A* CH. Surprisingly, besides age, we did not find any significant association between certain traditional risk factors for atherosclerosis, such as a history of smoking, hypertension, T2D, BMI, and LDL-C, and the risk for incident CH, contrary to previous cross-sectional observations related to prevalent CH. Our findings aligned with a recent longitudinal study demonstrating no association between smoking, being overweight, and acquiring new CH mutations^26^. Another study reported an inverse association between HDL-C and clonal expansion^28^. While we observed a nominal association between non-HDL-C and incident *TET2* CH in univariable analysis, this association did not hold in a fully adjusted logistic regression model.

Our genetic analyses provide compelling evidence for a shared genetic basis between prevalent and incident CH. For the first time, we demonstrated strong associations between prevalent CH PRS and increased odds of incident CH, underscoring the predictive value of CH PRS in identifying individuals at risk of developing CH. Furthermore, the genetic variants previously linked to prevalent CH^13^ also showed associations with incident CH in our study using the ARIC dataset. Specifically, we observed significant associations for *SMC4*, *TERT*, *ATM*, *TCL1A*, and *SETBP1* loci for incident CH. These findings further support the notion of a shared genetic underpinning between prevalent and incident CH, emphasizing the relevance of these specific loci in the development and progression of clonal hematopoiesis. Importantly, these results provide evidence for the causality of key genes in the development of CH itself.

However, it is worth noting that only a quarter of the prevalent CH-associated loci were associated with incident CH. Notably, certain loci, including *PARP1*, *LY75*, *SENP7*, *TET2*, *CD164*, and *ITPR2*, showed no association with incident CH. This could be due to lower statistical power, poor imputation quality, or true biological differences between prevalent and incident CH. The *CD164* locus is an example of the latter, where this locus is strongly associated with prevalent CH^13–15^ but lacks any association with incident CH. The opposite effect directions observed at the *CD164* locus for incident *DNMT3A* and *TET2* CH could explain this null association in overall CH. Previously we and others reported opposite associations at the *TCL1A* locus with prevalent *DNMT3A* and *TET2* CH, leading to a null association with overall CH^1,13–15^. This study found an association at the *TCL1A* locus with incident overall CH and *TET2* CH but not with incident *DNMT3A* CH. Our findings align with recent research demonstrating the involvement of *TCL1A* in the expansion of various non-*DNMT3A* CH subtypes, including *TET2*.^6^

The ongoing debate surrounding the relationship between CH and the development of ASCVD has led to investigations exploring the role of CH if it triggers inflammation and contributes to the development of ASCVD^4,29^, or if ASCVD itself or clinical risk factors for ASCVD promotes the development of CH^30^, as well as a potential bidirectional link between CH and inflammation^31^. While baseline ASCVD status showed a nominal association with incident *DNMT3A* and *TET2* CH, the associations were not statistically significant and were in opposite directions. Secondary analyses revealed a strong link between dyslipidemia and incident *TET2* and potentially *ASXL1* CH, while no such associations were found for *DNMT3A* or overall CH. These findings partially support the hypothesis that dyslipidemia may influence CH development^30^.

Nonetheless, explanations based solely on ASCVD (or associated risk factors) are insufficient in understanding the development of CH among older adults. The results from our clinical and genetic analyses point to intricate relationships between environmental and genetic risk factors in the development of clonal hematopoiesis, providing strong evidence for a bidirectional association between CH and inflammation. Notably, studies in mice and zebrafish support a positive feedback loop between CH expansion and inflammation, where CH promotes inflammation, and the relative fitness and resistance to inflammation of CH clones further fuel clonal expansion, creating a vicious cycle of inflammation and expansion^31–33^. These findings highlight the complexity of the interactions between exposome, inherited, and somatic genomes, shedding light on the pathogenesis of CH in the context of (inflamm-)aging^34–36^. Further research is crucial to fully unravel these intricate relationships’ underlying mechanisms and potential therapeutic implications.

Although our study demonstrates associations between environmental and genetic determinants, important limitations should be considered. First, variability in CH ascertainment: CH was determined using two different sequencing platforms. The technical differences, albeit with good correlation, precluded the ability to compare clonal trajectories and growth rates and investigate factors associated with VAF changes but were suitable for incident CH analyses. Second, homogeneity in follow-up time: the study was constrained by a lack of heterogeneity in the follow-up time among participants as well as a single follow-up sampling. This limited the ability to perform time-to-event analyses between the baseline exposures and the incidence of CH. Longer follow-up durations and varying time intervals could provide a more comprehensive understanding of the relationship between exposures and the development of CH. Third, although the long duration of almost two decades between time points was a strength in regards to examining the impact of age, a substantial number of individuals died during this time period and as shown in **Suppl. Table 1**. Participants without follow-up WES had more clinical ASCVD, T2D, hypertension, cholesterol medication usage, increased BMI, and unfavorable lipid profiles. Thus, individuals with the most severe ASCVD and perhaps the greatest baseline inflammation were more likely to die before the follow-up visit (ARIC visit 5), limiting our power and biasing our results towards the null. Fourth, the study may have suffered from reduced statistical power, which could have influenced the observed associations between exposures and outcomes. Larger sample sizes would enhance the study’s statistical power and provide more reliable conclusions. Therefore, these limitations should be considered when interpreting the findings, and future studies should aim to address these issues to further advance our understanding of the relationships between environmental and genetic determinants and their impact on clonal hematopoiesis.

Our comprehensive long-term longitudinal assessment provides new insights into the factors that promote incident CH in older adults. We find that age at baseline, sex, and dyslipidemia are significant predictors of incident CH, while ASCVD and traditional risk factors for ASCVD are not. We also find that the factors driving clinically relevant clonal expansion (VAF ≥ 2%) may vary, to some extent, among different CH driver genes. Additionally, our research demonstrates a shared genetic basis between prevalent and incident CH, further supporting the notion that CH can evolve independently of prevalent ASCVD and traditional ASCVD risk factors. Taken together, these results support a bidirectional relationship between CH and inflammation. Early identification of incident CH presents an opportunity for timely intervention and preventive measures. These findings underscore the importance of further research to better understand the mechanisms underlying incident CH and to develop strategies for its early detection and effective management.

## Methods

### Study samples

There were 10,881 ARIC participants with WES data at baseline sequenced using the HiSeq 2000 platform (Illumina, Inc., CA) (ARIC sub-study phs000668 CHARGE-S^37^https://www.ncbi.nlm.nih.gov/projects/gap/cgi-bin/study.cgi?study_id=phs000668.v6.p2). Baseline samples were from five visits with age ranges 44-84 years (Mean=57.3; Median=57.0; SD=6.06). WES was also generated using the NovaSeq 6000 platform (Illumina, Inc., CA) from 4,233 longitudinal visit samples (ARIC Visit 05). Among the longitudinal visit samples, 4,187 without hematologic malignancy had baseline WES data (detail study design in **Suppl. Fig. 1**).

### Detection of clonal hematopoiesis (CH)

Somatic mutations were identified from WES using Mutect2 software^38^ in the Terra platform (https://portal.firecloud.org/?return=terra#methods/gatk/mutect2-gatk4/21), and CH was detected using a publicly available pipeline (https://app.terra.bio/#workspaces/terra-outreach/CHIP-Detection-Mutect2/). To minimize potential artifacts in the Mutect2 calls, a panel-of-normal (PON) was created from 100 random HiSeq WES from the youngest participants, while 1000 Genomes PON was used for the NovaSeq WES. Besides, Genome Aggregation Database (gnomAD) was used to limit germline variants in the somatic mutation call. Mutect2 calls were further filtered, and variants were kept if (i) total depth of coverage ≥20, (ii) number of reads supporting the alternate allele ≥3, (iii) ≥1 read in both forward and reverse direction supporting the alternate allele, (iv) variant allele fraction ≥2%, (v) gnomAD allele frequency ≤0.001 (not hotspot mutations). Finally, CH mutations that passed sequence-based filtering were manually curated. Pathogenic variants were queried in 69 genes known to drive clonal hematopoiesis and myeloid malignancies^4,39^ to identify CH. The detailed CHIP calling pipeline was previously reported by ^1,15^ (https://app.terra.bio/#workspaces/terra-outreach/CHIP-Detection-Mutect).

### Hotspot mutations in *U2AF1*

A special approach was used to identify somatic variants in *U2AF1* since an erroneous segmental duplication in the hg38 reference genome resulted in a mapping score of zero for this gene during the sequence alignment from FASTQ to BAM/CRAM. We used a custom script (https://github.com/MMesbahU/U2AF1_pileup) to recover hotspot mutations: S34F, S34Y, R156H, Q157P, and Q157R. A minimum of 5 supporting reads for alternate alleles was required to include a somatic mutation in *U2AF1*.

### HiSeq vs. NovaSeq WES

To analyze the concordance between the two platforms used for WES, we re-sequenced 786 baseline samples using the NovaSeq 6000 platform (Illumina, Inc., CA). CH was detected using the same pipeline described above. We compared CH status “yes/no” at VAF≥2% and VAF≥10%. We also performed Pearson’s correlation between the VAF estimates when the same clone was detected from WES generated by the two platforms.

### Associations between baseline risk factors and incident CH

Incident CH was defined as the presence of a clone with a VAF of ≥2% at the follow-up visit in individuals who did not have a prevalent CH clone (or CH at VAF<2%) at baseline. The CH mutations were considered if they met the following criteria: a read depth ≥20, a minimum of 3 supporting reads for the alternative allele, and at least one read from both the forward and reverse directions.

Univariable and multivariable analyses were performed using a logistic regression model to examine the associations between incident CH and baseline risk factors. The tested risk factors included age, sex, race, smoking status, body mass index (BMI), high-density lipoprotein cholesterol (HDL-C), non-HDL-C, disease status for type 2 diabetes (T2D), atherosclerotic cardiovascular disease (ASCVD, including ischemic stroke and coronary heart disease), and hypertension. Additional covariates considered in the analysis included cholesterol medication usage, baseline visits, and visit centers.

Secondary analyses were performed to test the association between exposures such as smoking categories (never vs former or current smoker), BMI categories (BMI<25 vs. 25≤BMI≤30 or BMI>30 kg/m^2^), triglyceride to HDL-C (TG/HDL-C) ratio and dyslipidemia, and incident CH. Interactions between sex and smoking status were also tested in a fully adjusted model for association with incident CH.

Here, dyslipidemia was defined as a binary variable (“yes/no”) based on the following criteria: individuals with total cholesterol ≥240, triglyceride ≥200, LDL-C ≥160, HDL-C <40 in men or HDL-C <50 mg/dl in women, and/or individuals on statin therapy. Inverse rank normalization was performed before the analysis to account for potential variations in the distribution of BMI, HDL-C, non-HDL-C, and TG/HDL-C values. To account for multiple comparisons in the analysis, a significance threshold of <0.0025 was employed, considering 20 independent tests at a 5% significance level.

### Single-variant association analysis

Imputed genotype data from the ARIC sub-study GENEVA (dbGaP accession phs000090 “phg000248” https://www.ncbi.nlm.nih.gov/projects/gap/cgi-bin/study.cgi?study_id=phs000090.v8.p2) was used for genetic analyses. From dbGaP, we downloaded genotype data (Affymetrix 6.0 SNP array) imputed to the whole-genome sequence using the 1000 Genomes reference panel (www.1000genomes.org, June 2011 release)^40^ using IMUTE2 software^41^ (details in dbGaP phs000090/phg000248/). Imputed GWAS data and WES were available for 637 AA participants and 2,377 EA participants. Variants with a minor allele frequency >1%, imputation accuracy (INFO score) ≥0.30, and significant association (*P*<5.0E-8) with prevalent CH in the UK Biobank^13^ were considered for association analysis. We performed ancestry-stratified single-variant associations, adjusted for age, age^2^, sex, top ten principal components of ancestry, and batch effect, followed by multi-ancestry inverse-variance weighted fixed-effect meta-analysis for incident overall CH, *DNMT3A*, and *TET2* CH. REGENIE software^42^ was used for single-variant associations, and GWAMA software^43^ for multi-ancestry metanalysis (scripts available in the Code section). Variants with an association *P*<0.05 were considered significant for these analyses.

### Prevalent CH polygenic risk score (PRS)

Prevalent CH PRS were derived for 637 AA and 2,376 EA participants using weights of 21 single-nucleotide variants that were conditionally independent and significantly associated with prevalence CH in the UK Biobank at *P*<5E-8 ^13^. Effects for the risk-increasing allele were considered and PLINK (version v2.00a3LM) “score” function was used to derive prevalent CH PRS. PRS was standardized to have a mean of zero and SD of 1. We performed associations for prevent CH PRS with incident CH categories in ARIC AA and EA participants using multivariable logistic regressions adjusted for age, sex, smoking status (never vs. ever), top five principal components of ancestry, and batch effect, followed by inverse-variance weighted fixed-effect multi-ancestry meta-analyses (scripts available in the Code section).

### Use of large language models

Advanced language models like Grammarly, Inc., Bard (https://bard.google.com/), and ChatGPT (version May 24; https://chat.openai.com/) were used to enhance grammatical accuracy and improve sentence structure and clarity.

## Code availability

The complete scripts used in this study will be available at https://github.com/MMesbahU/longitudinal-profiling-of-clonal-hematopoiesis.

## Data Availability

Individual-level phenotypes and whole-exome sequencing data from the ARIC baseline visits participants are available in dbGaP (https://www.ncbi.nlm.nih.gov/gap/) accession code phs000668. Imputed genotype data from the ARIC sub-study GENEVA is available in dbGaP (accession phs000090 phg000248 https://www.ncbi.nlm.nih.gov/projects/gap/cgi-bin/study.cgi?study_id=phs000090.v8.p2). ARIC Visit-5 WES and individual-level data are available via controlled access ancillary study proposal. Timeline for the approval process range from 3-6 weeks for ARIC ancillary studies, with specific criteria and proposal forms available at https://sites.cscc.unc.edu/aric/ancillary-studies-pfg. Codes used in this study are available on GitHub.

## Acknowledgment

The Atherosclerosis Risk in Communities study has been funded in whole or in part with Federal funds from the National Heart, Lung, and Blood Institute, National Institutes of Health, Department of Health and Human Services, under Contract nos. (HHSN268201700001I, HHSN268201700002I, HHSN268201700003I, HHSN268201700004I, HHSN268201700005I).

Funding was also supported by R01HL087641, R01HL059367 and R01HL086694; National Human Genome Research Institute contract U01HG004402; and National Institutes of Health contract HHSN268200625226C. Infrastructure was partly supported by Grant Number UL1RR025005, a component of the National Institutes of Health and NIH Roadmap for Medical Research. Funding support for “Building on GWAS for NHLBI-diseases: the U.S. CHARGE consortium” was provided by the NIH through the American Recovery and Reinvestment Act of 2009 (ARRA) (5RC2HL102419). CHARGE sequencing was carried out at the Baylor College of Medicine Human Genome Sequencing Center (U54 HG003273 and R01HL086694). Funding for GO ESP was provided by NHLBI grants RC2 HL-103010 (HeartGO) and exome sequencing was performed through NHLBI grants RC2 HL-102925 (BroadGO) and RC2 HL-102926 (SeattleGO). The authors thank the staff and participants of the ARIC study for their important contributions. The authors also thank Mrs. Leslie Gaffney from the Broad Research Communication Lab for her valuable assistance in improving the display items.

