## Supplemental Table1 and Figures 1-6 for "Long-term longitudinal analysis of 4,187 participants reveals new insights into determinants of incident clonal hematopoiesis"

### Supplementary materials

#### Supplementary Tables

**Supplementary Table 1. Baseline characteristics of the Atherosclerosis Risk in Communities Study (N=10,871)**

| Characteristics | | Without Follow-up WES | With Follow-up WES | *P* |
|---|---|---|---|---|
| **N** | | 6,684 | 4,187 | - |
| **Age, mean (SD), y** | | 58.5 (6.1) | 55.5 (5.5) | 3.0E-149 |
| **Sex** | **Female, n (%)** | 3,664 (54.8) | 2,478 (59.2) | 8.6E-6* |
| | **Male, n (%)** | 3,020 (45.2) | 1,709 (40.8) | |
| **Ancestry** | **African American (AA)** | 2,171 (32.5) | 951 (22.7) | 8.2E-28* |
| | **European American (EA)** | 4,513 (67.5) | 3,236 (77.3) | |
| **Ever Smoker, n (%)** | | 4,212 (63.2) | 2,269 (54.3) | 5.0E-20 |
| **BMI, mean (SD), kg/m$^2$** | | 28.5 (5.8) | 27.8 (5.1) | 9.6E-12 |
| **SBP, mean (SD), mmHg** | | 124.8 (19.5) | 118.4 (16.3) | 2.5E-74 |
| **DBP, mean (SD), mmHg** | | 72.7 (10.8) | 72.4 (9.8) | 0.13 |
| **Cholesterol medication, n (%)** | | 470 (7.1) | 231 (5.5) | 0.001 |
| **Hypertension, n (%)** | | 2,879 (43.3) | 1,194 (28.6) | 4.8E-56 |
| **T2D, n (%)** | | 1,309 (19.7) | 349 (8.4) | 3.9E-67 |
| **CHD, n (%)** | | 544 (8.1) | 127 (3.0) | 8.9E-33 |
| **IS, n (%)** | | 215 (3.3) | 42 (1.03) | 7.1E-17 |
| **HF, n (%)** | | 438 (6.7) | 113 (2.7) | 1.0E-22 |
| **Total cholesterol, mean (SD), mg/dl** | | 211.1 (40.4) | 208.1 (36.5) | 4.7E-5 |
| **LDL-C, mean (SD), mg/dl** | | 134.0 (36.8) | 130.6 (34.4) | 1.4E-6 |
| **HDL-C, mean (SD), mg/dl** | | 49.8 (17) | 52 (17) | 2.8E-11 |
| **Triglycerides, median (IQR), mg/dl** | | 116.0 (84) | 110.0 (74) | 2.3E-8# |
| **$CH_{\geq 2\%\ VAF}$ prevalence, n (%)** | | 635 (9.5) | 457 (10.9) | 0.019 |
| **$CH_{\geq 10\%\ VAF}$ prevalence, n (%)** | | 287 (4.3) | 161 (3.8) | 0.25 |
| **Follow-up, mean (range), y** | | - | 20.3 (5-27) | - |

*P*-values are from a two-sided *t-test* comparing baseline samples with and without prevalent CH. **P* from Pearson's *Chi-squared* test. #*P* from a two-sided Wilcoxon rank sum test. BMI: Body mass index; CH: clonal hematopoiesis; CHD: Coronary heart disease; DBP: Diastolic blood pressure; HDL-C: high-density lipoprotein cholesterol; HF: Heart failure; IQR: interquartile range; IS: ischemic stroke; LDL-C: low-density lipoprotein cholesterol; SBP: Systolic blood pressure; SD: standard deviation; T2D: Type 2 Diabetes; VAF: variant allele fraction; WES: whole-exome sequence.

**Supplementary Table 2. List of clonal hematopoiesis mutations detected in the baseline visit samples.**

**Supplementary Table 3. List of clonal hematopoiesis mutations detected in the follow-up visit samples.**

**Supplementary Table 4. Conditionally independent genome-wide significant variants used for deriving prevalent CH polygenic risk score.** This table lists 21 conditionally independent genome-wide significant variants from Kessler, et al. [13]. These variants were used for deriving the polygenic risk score. REF: Reference Allele; ALT: Alternate Allele; AAF: Alternate Allele Frequency; COJO: Conditional and Joint analysis; RA: Risk Allele; RAF: Risk Allele Frequency.

**Supplementary Table 5. Multi-ancestry meta-analysis of single variant association summary statistics for incident overall CH in the Atherosclerosis Risk in Communities (ARIC) Study.** Significant variants ($P$<5.0E-8) from prevalent CH loci from Kessler, et al. [13] were considered for the association tests. $P$<0.05 in the ARIC study highlighted in bold.

**Supplementary Table 6. Multi-ancestry meta-analysis of single variant association summary statistics for incident *DNMT3A* CH in the Atherosclerosis Risk in Communities (ARIC) Study.** Significant variants ($P$<5.0E-8) from prevalent CH loci from Kessler, et al. [13] were considered for the association tests. $P$<0.05 in the ARIC study highlighted in bold.

**Supplementary Table 7. Multi-ancestry meta-analysis of single variant association summary statistics for incident *TET2* CH in the Atherosclerosis Risk in Communities (ARIC) Study.** Significant variants ($P$<5.0E-8) from prevalent CH loci from Kessler, et al. [13] were considered for the association tests. $P$<0.05 in the ARIC study highlighted in bold.

#### Supplementary Figures

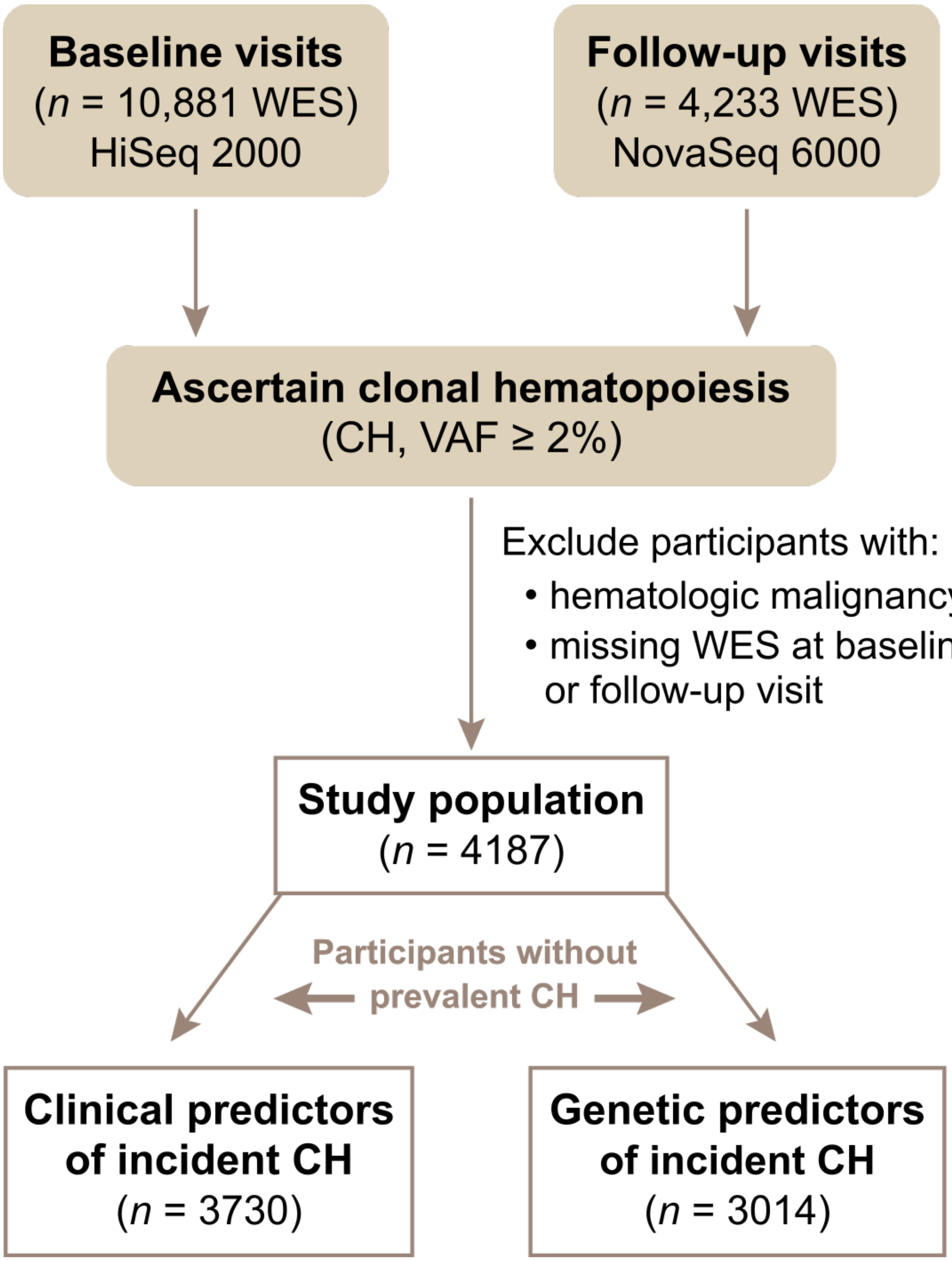


**Supplementary Fig. 1 | Flow diagram of the study design.** HiSeq and NovaSeq are two Illumina sequencing platforms used for WES in the ARIC Study. CH: clonal hematopoiesis; WES: whole-exome sequence.

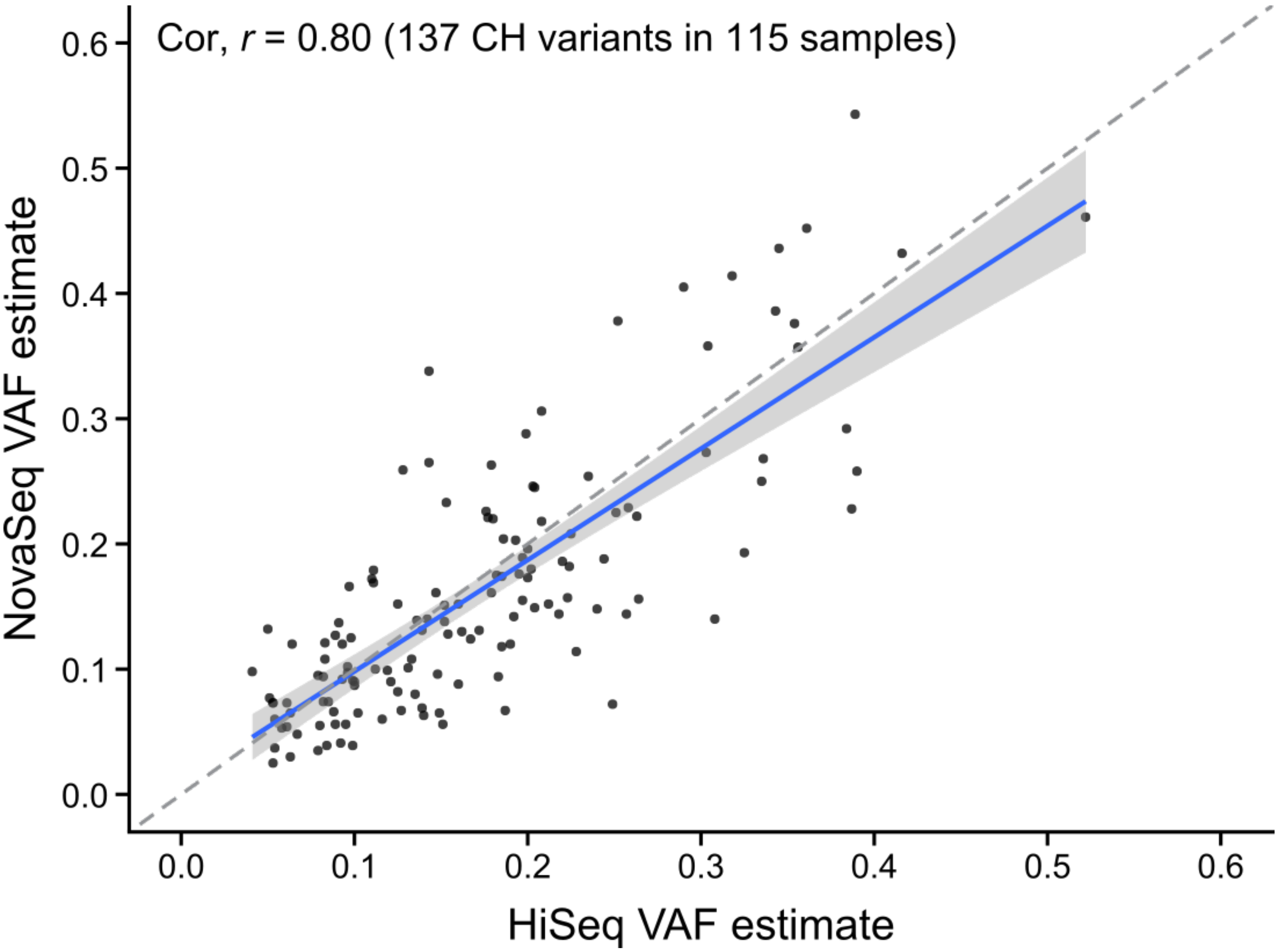


**Supplementary Fig. 2 | Estimates of variant allele fraction (VAF) from the HiSeq 2000 vs. NovaSeq 6000 sequencing platforms. In the Atherosclerosis Risk in Communities Study,** Illumina sequencing platform Hiseq 2000 was used for whole-exome sequencing (WES) for the baseline samples, and NovaSeq 6000 was used for longitudinal visit samples. To estimate the correlation of VAF and concordance of CH ascertainment using the WES from both platforms, repeat sequencing using NovaSeq was performed for 786 baseline visit samples. Here, each dot represents a CH mutation detected by both sequencing platforms. Overall Pearson's correlation between HiSeq and NovaSeq VAF is $r$=0.80. CH: clonal hematopoiesis.

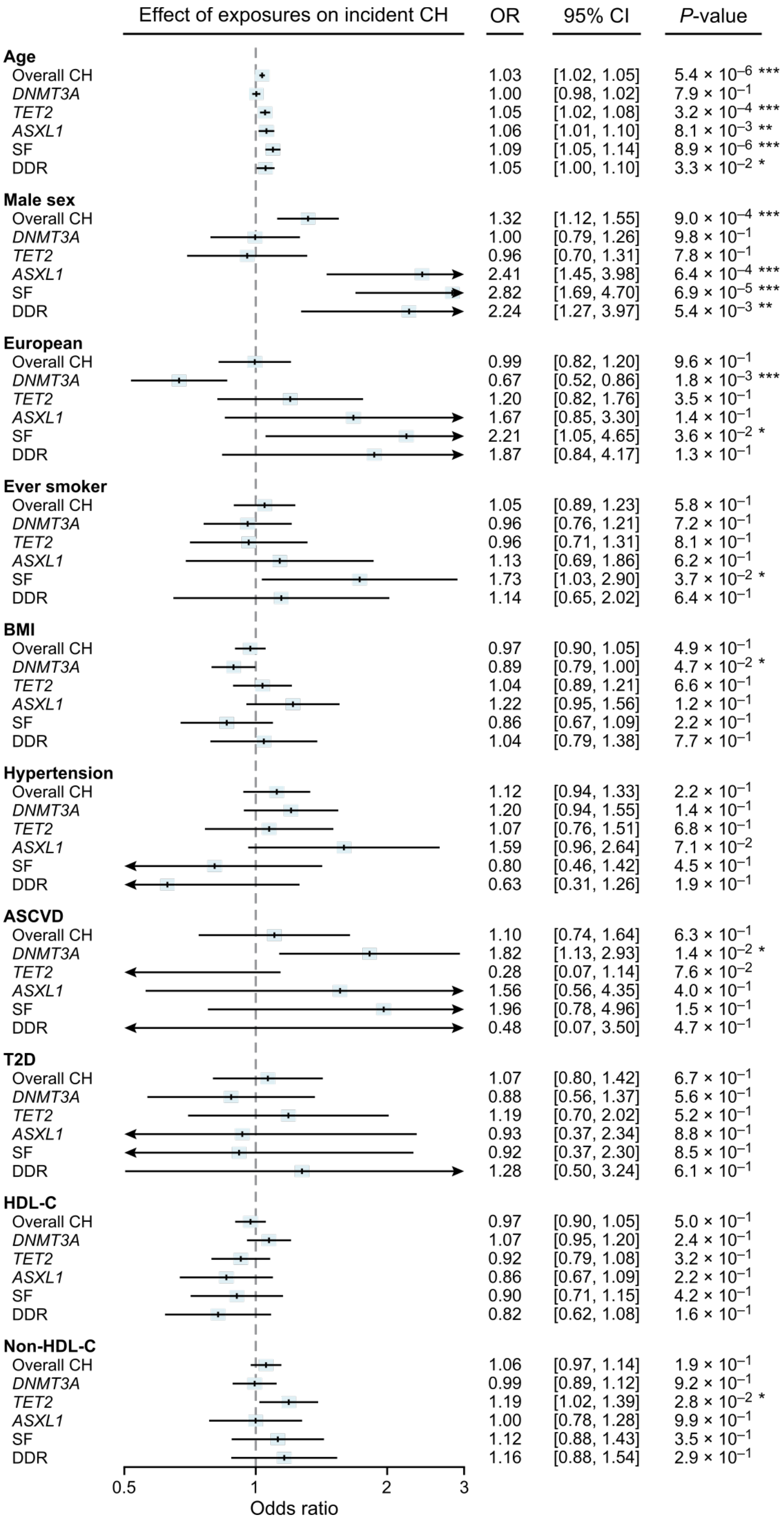
Effect of exposures on incident CH
OR
95% CI
*P*-value
**Age**
Overall CH 1.03 [1.02, 1.05] $5.4 \times 10^{-6}$ ***
*DNMT3A* 1.00 [0.98, 1.02] $7.9 \times 10^{-1}$
*TET2* 1.05 [1.02, 1.08] $3.2 \times 10^{-4}$ ***
*ASXL1* 1.06 [1.01, 1.10] $8.1 \times 10^{-3}$ **
SF 1.09 [1.05, 1.14] $8.9 \times 10^{-6}$ ***
DDR 1.05 [1.00, 1.10] $3.3 \times 10^{-2}$ *
**Male sex**
Overall CH 1.32 [1.12, 1.55] $9.0 \times 10^{-4}$ ***
*DNMT3A* 1.00 [0.79, 1.26] $9.8 \times 10^{-1}$
*TET2* 0.96 [0.70, 1.31] $7.8 \times 10^{-1}$
*ASXL1* 2.41 [1.45, 3.98] $6.4 \times 10^{-4}$ ***
SF 2.82 [1.69, 4.70] $6.9 \times 10^{-5}$ ***
DDR 2.24 [1.27, 3.97] $5.4 \times 10^{-3}$ **
**European**
Overall CH 0.99 [0.82, 1.20] $9.6 \times 10^{-1}$
*DNMT3A* 0.67 [0.52, 0.86] $1.8 \times 10^{-3}$ ***
*TET2* 1.20 [0.82, 1.76] $3.5 \times 10^{-1}$
*ASXL1* 1.67 [0.85, 3.30] $1.4 \times 10^{-1}$
SF 2.21 [1.05, 4.65] $3.6 \times 10^{-2}$ *
DDR 1.87 [0.84, 4.17] $1.3 \times 10^{-1}$
**Ever smoker**
Overall CH 1.05 [0.89, 1.23] $5.8 \times 10^{-1}$
*DNMT3A* 0.96 [0.76, 1.21] $7.2 \times 10^{-1}$
*TET2* 0.96 [0.71, 1.31] $8.1 \times 10^{-1}$
*ASXL1* 1.13 [0.69, 1.86] $6.2 \times 10^{-1}$
SF 1.73 [1.03, 2.90] $3.7 \times 10^{-2}$ *
DDR 1.14 [0.65, 2.02] $6.4 \times 10^{-1}$
**BMI**
Overall CH 0.97 [0.90, 1.05] $4.9 \times 10^{-1}$
*DNMT3A* 0.89 [0.79, 1.00] $4.7 \times 10^{-2}$ *
*TET2* 1.04 [0.89, 1.21] $6.6 \times 10^{-1}$
*ASXL1* 1.22 [0.95, 1.56] $1.2 \times 10^{-1}$
SF 0.86 [0.67, 1.09] $2.2 \times 10^{-1}$
DDR 1.04 [0.79, 1.38] $7.7 \times 10^{-1}$
**Hypertension**
Overall CH 1.12 [0.94, 1.33] $2.2 \times 10^{-1}$
*DNMT3A* 1.20 [0.94, 1.55] $1.4 \times 10^{-1}$
*TET2* 1.07 [0.76, 1.51] $6.8 \times 10^{-1}$
*ASXL1* 1.59 [0.96, 2.64] $7.1 \times 10^{-2}$
SF 0.80 [0.46, 1.42] $4.5 \times 10^{-1}$
DDR 0.63 [0.31, 1.26] $1.9 \times 10^{-1}$
**ASCVD**
Overall CH 1.10 [0.74, 1.64] $6.3 \times 10^{-1}$
*DNMT3A* 1.82 [1.13, 2.93] $1.4 \times 10^{-2}$ *
*TET2* 0.28 [0.07, 1.14] $7.6 \times 10^{-2}$
*ASXL1* 1.56 [0.56, 4.35] $4.0 \times 10^{-1}$
SF 1.96 [0.78, 4.96] $1.5 \times 10^{-1}$
DDR 0.48 [0.07, 3.50] $4.7 \times 10^{-1}$
**T2D**
Overall CH 1.07 [0.80, 1.42] $6.7 \times 10^{-1}$
*DNMT3A* 0.88 [0.56, 1.37] $5.6 \times 10^{-1}$
*TET2* 1.19 [0.70, 2.02] $5.2 \times 10^{-1}$
*ASXL1* 0.93 [0.37, 2.34] $8.8 \times 10^{-1}$
SF 0.92 [0.37, 2.30] $8.5 \times 10^{-1}$
DDR 1.28 [0.50, 3.24] $6.1 \times 10^{-1}$
**HDL-C**
Overall CH 0.97 [0.90, 1.05] $5.0 \times 10^{-1}$
*DNMT3A* 1.07 [0.95, 1.20] $2.4 \times 10^{-1}$
*TET2* 0.92 [0.79, 1.08] $3.2 \times 10^{-1}$
*ASXL1* 0.86 [0.67, 1.09] $2.2 \times 10^{-1}$
SF 0.90 [0.71, 1.15] $4.2 \times 10^{-1}$
DDR 0.82 [0.62, 1.08] $1.6 \times 10^{-1}$
**Non-HDL-C**
Overall CH 1.06 [0.97, 1.14] $1.9 \times 10^{-1}$
*DNMT3A* 0.99 [0.89, 1.12] $9.2 \times 10^{-1}$
*TET2* 1.19 [1.02, 1.39] $2.8 \times 10^{-2}$ *
*ASXL1* 1.00 [0.78, 1.28] $9.9 \times 10^{-1}$
SF 1.12 [0.88, 1.43] $3.5 \times 10^{-1}$
DDR 1.16 [0.88, 1.54] $2.9 \times 10^{-1}$
0.5
1
2
3
Odds ratio

**Supplementary Fig. 3 | Univariable association (logistic regression) between baseline risk factors and incident CH.** BMI, HDL-C, and non-HDL-C values were transformed by inverse rank normalization. ASCVD: atherosclerotic cardiovascular disease (including coronary heart disease or ischemic stroke); BMI: body mass index; DDR: DNA damage response genes (*PPM1D* or *TP53*); CH: clonal hematopoiesis; HDL-C: high-density lipoprotein cholesterol; SF: CH in splicing factors genes (*SF3B1*, *U2AF1*, *SRSF2* or *ZRSR2*); T2D: type 2 diabetes. ***: $P$<0.0025 (0.05/20); **: $P$<0.01; *: $P$<0.05.

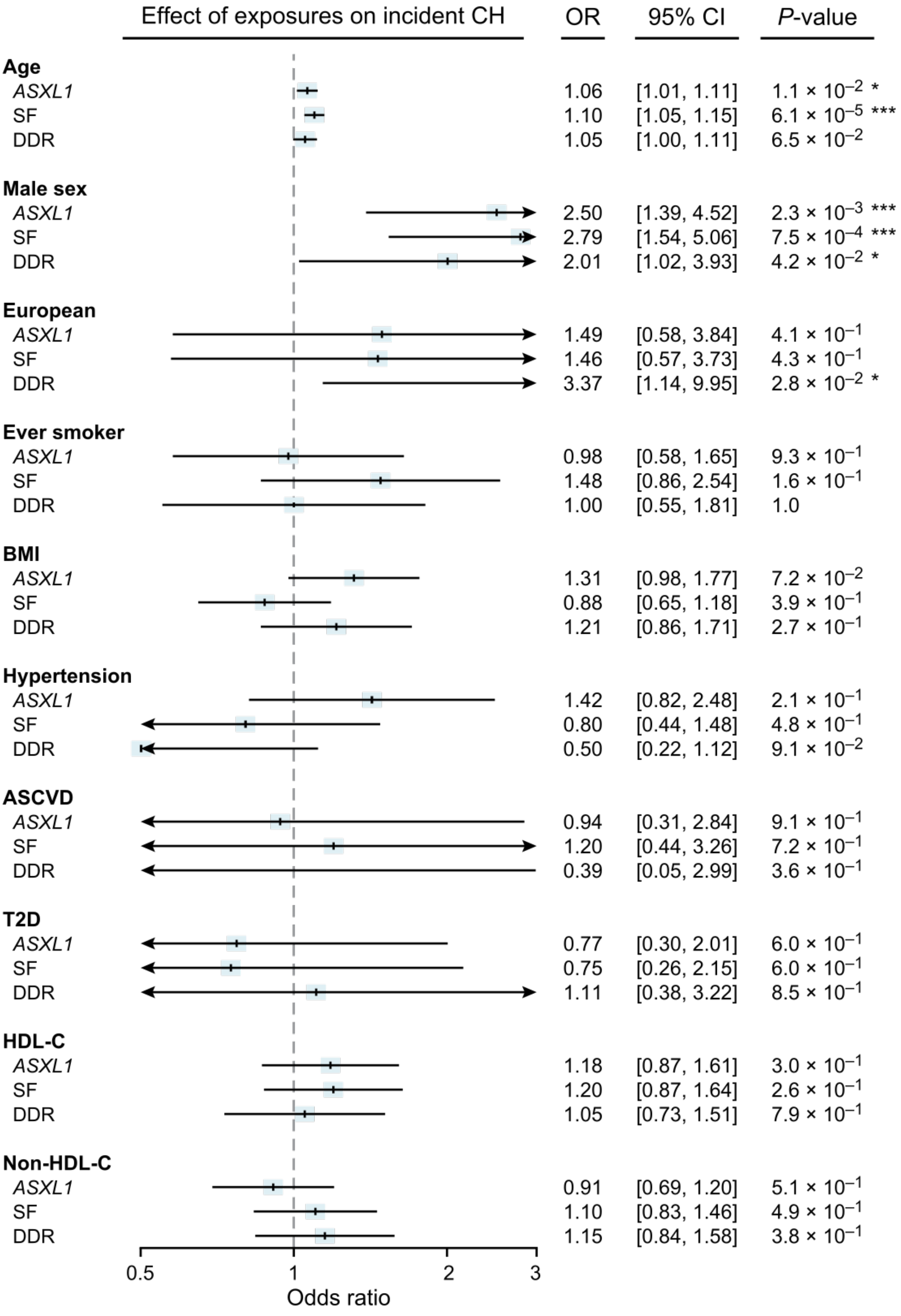

Effect of exposures on incident CH
OR
95% CI
P-value
Age
ASXL1 1.06 [1.01, 1.11] 1.1 × 10−2 *
SF 1.10 [1.05, 1.15] 6.1 × 10−5 ***
DDR 1.05 [1.00, 1.11] 6.5 × 10−2
Male sex
ASXL1 2.50 [1.39, 4.52] 2.3 × 10−3 ***
SF 2.79 [1.54, 5.06] 7.5 × 10−4 ***
DDR 2.01 [1.02, 3.93] 4.2 × 10−2 *
European
ASXL1 1.49 [0.58, 3.84] 4.1 × 10−1
SF 1.46 [0.57, 3.73] 4.3 × 10−1
DDR 3.37 [1.14, 9.95] 2.8 × 10−2 *
Ever smoker
ASXL1 0.98 [0.58, 1.65] 9.3 × 10−1
SF 1.48 [0.86, 2.54] 1.6 × 10−1
DDR 1.00 [0.55, 1.81] 1.0
BMI
ASXL1 1.31 [0.98, 1.77] 7.2 × 10−2
SF 0.88 [0.65, 1.18] 3.9 × 10−1
DDR 1.21 [0.86, 1.71] 2.7 × 10−1
Hypertension
ASXL1 1.42 [0.82, 2.48] 2.1 × 10−1
SF 0.80 [0.44, 1.48] 4.8 × 10−1
DDR 0.50 [0.22, 1.12] 9.1 × 10−2
ASCVD
ASXL1 0.94 [0.31, 2.84] 9.1 × 10−1
SF 1.20 [0.44, 3.26] 7.2 × 10−1
DDR 0.39 [0.05, 2.99] 3.6 × 10−1
T2D
ASXL1 0.77 [0.30, 2.01] 6.0 × 10−1
SF 0.75 [0.26, 2.15] 6.0 × 10−1
DDR 1.11 [0.38, 3.22] 8.5 × 10−1
HDL-C
ASXL1 1.18 [0.87, 1.61] 3.0 × 10−1
SF 1.20 [0.87, 1.64] 2.6 × 10−1
DDR 1.05 [0.73, 1.51] 7.9 × 10−1
Non-HDL-C
ASXL1 0.91 [0.69, 1.20] 5.1 × 10−1
SF 1.10 [0.83, 1.46] 4.9 × 10−1
DDR 1.15 [0.84, 1.58] 3.8 × 10−1
0.5
1
2
3
Odds ratio

**Supplementary Fig. 4 | Multivariable adjusted logistic regression for incident *ASXL1*, SF, and DDR CH vs baseline risk factors.** Multivariable-adjusted logistic regression analyses examined the relationship between incident *ASXL1*, SF, and DDR CH and baseline risk factors. The logistic regression model included several covariates: age, sex, race, BMI, HDL-C, non-HDL-C, cholesterol medication usage, history of smoking, hypertension, ASCVD, T2D, baseline visits, and visits center. Inverse rank normalized BMI, HDL-C, and non-HDL-C values were used in the analyses. The results show significant associations between age at enrollment and male sex and incident CH. ASCVD: atherosclerotic cardiovascular disease (including coronary heart disease or ischemic stroke); BMI: body mass index; CH: clonal hematopoiesis; DDR: DNA damage response genes (*PPM1D* or *TP53*); HDL-C: high-density lipoprotein cholesterol; SF: CH in splicing factors genes (*SF3B1*, *U2AF1*, *SRSF2* or *ZRSR2*); T2D: type 2 diabetes. ***: $P<0.0025$ (0.05/20); **: $P<0.01$; *: $P<0.05$.

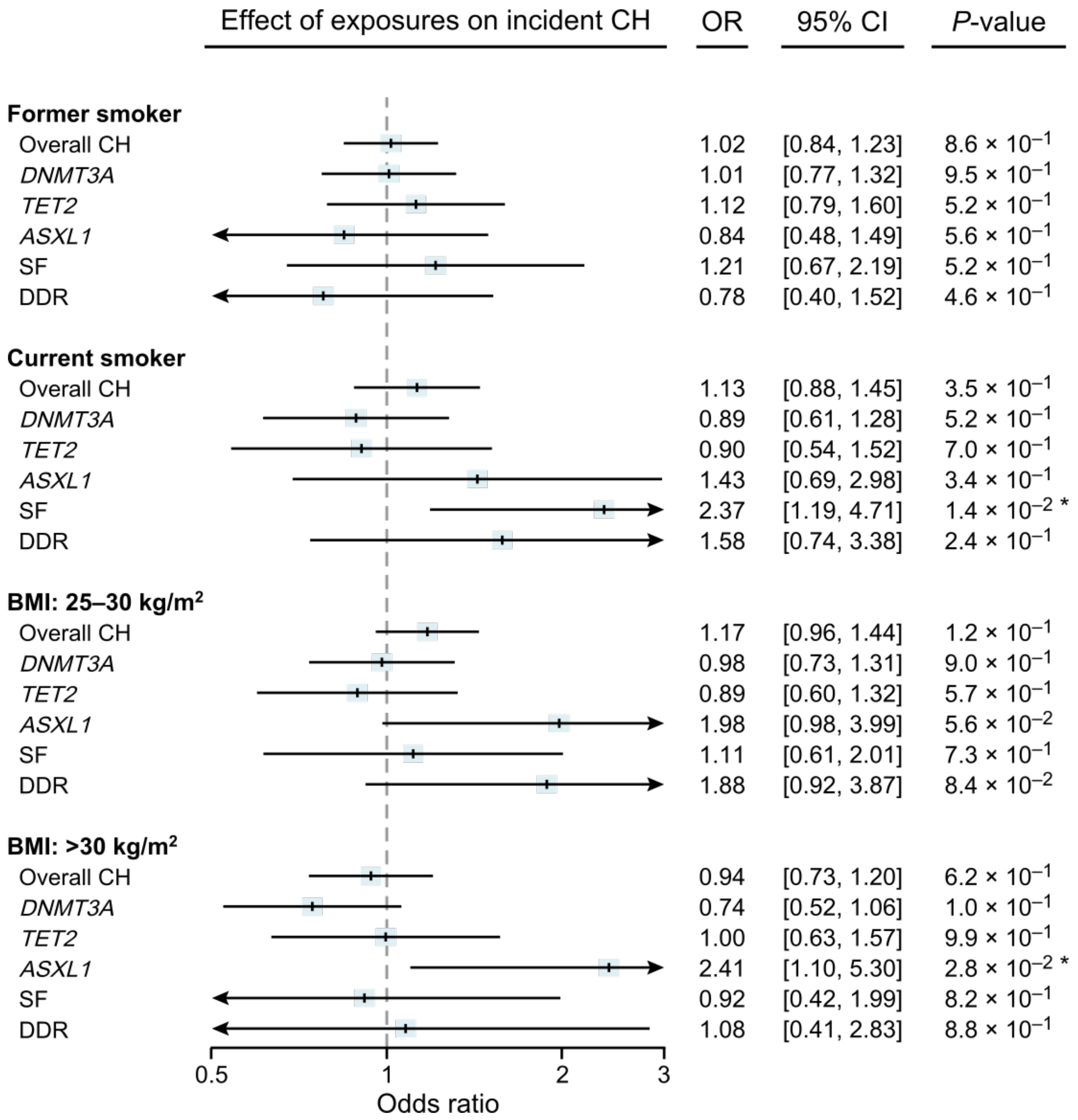


**Supplementary Fig. 5 | Logistic regression for incident CH vs smoking status (never vs former/current) and BMI categories.** The models included age, sex, race, BMI categories (BMI <25 vs. 25-30 or >30 kg/$m^2$), smoking status (never vs. former or current smokers), history of type-2 diabetes, hypertension, atherosclerotic cardiovascular disease, high-density lipoprotein cholesterol (HDL-C), non-HDL-C, cholesterol medication, baseline visit, and visit center. BMI: body mass index; CH: clonal hematopoiesis; DDR: DNA damage response genes (*PPM1D* or *TP53*); SF: CH in splicing factors genes (*SF3B1*, *U2AF1*, *SRSF2* or *ZRSR2*). ***: $P$<0.0025 (0.05/20); **: $P$<0.01; *: $P$<0.05.

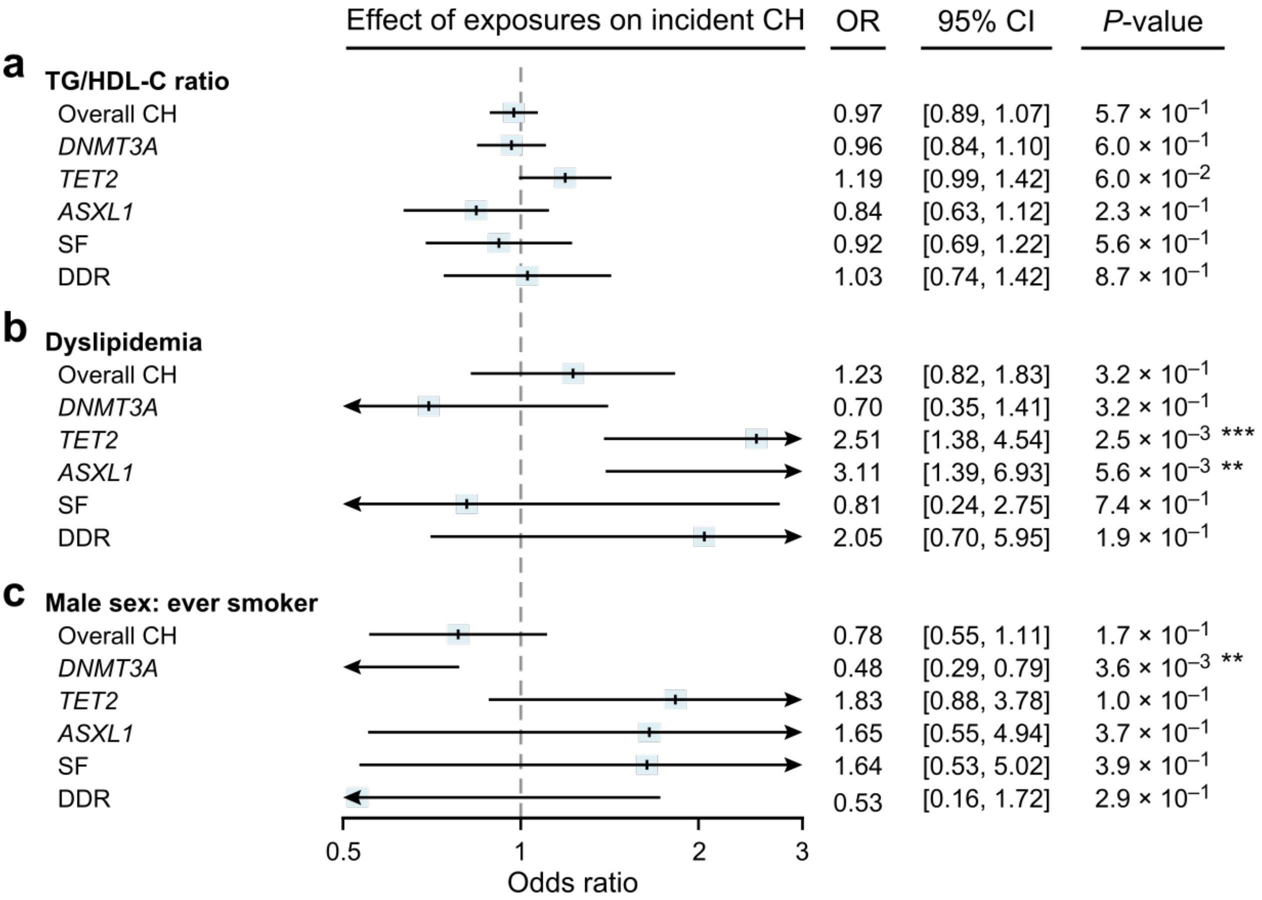


**Supplementary Fig. 6 | Associations between incident CH categories and (a) triglyceride to high-density lipoprotein cholesterol (TG/HDL-C) ratio, (b) dyslipidemia, and (c) male sex stratified by smoking status (never vs. ever) were examined.** The statistical models in (a-b) were adjusted for age, sex, race, smoking status, body mass index (BMI), history of type-2 diabetes, hypertension, atherosclerotic cardiovascular disease, baseline visit, and visit center. Additionally, in model (c), the analysis was further adjusted for HDL-C, non-HDL-C, and cholesterol medication usage. Inverse rank normalization was performed before the analysis to account for potential variations in the distribution of BMI, HDL-C, non-HDL-C, and TG/HDL-C values. Dyslipidemia was defined as "yes/no" based on the following criteria: individuals with total cholesterol ≥240, triglyceride ≥200, LDL-C ≥160, HDL-C <40 in men or HDL-C <50 mg/dL in women, and/or individuals on statin therapy. The results show a significant association between dyslipidemia and an increased odds of incident *TET2*, a nominal association between dyslipidemia and increased odds of incident *ASXL1,* and a nominal interaction between male sex and smoking status, indicating a lower risk for incident *DNMT3A*. CH: clonal hematopoiesis; DDR: DNA damage response genes (*PPM1D* or *TP53*); SF: CH in splicing factors genes (*SF3B1*, *U2AF1*, *SRSF2* or *ZRSR2*). ***: $P<0.0025$ (0.05/20); **: $P<0.01$; *: $P<0.05$.
